# A Network Analysis of HIV Civil Society Organizations to Enhance HIV Prevention Service Delivery in Blantyre, Malawi

**DOI:** 10.1101/2025.01.29.25321212

**Authors:** Rachel Haggard, Patricia Khomani, Yohane Kamgwira, Chimwemwe Mablekisi, Rosemary Ngaiyaye, Stanley Moyo, Dylan Green, Tyler Smith, Hannah Cooper, Charles B. Holmes, Sara Allinder, Deborah Hoege, Gift Kalawazira

**Affiliations:** Cooper/Smith, USA; Cooper/Smith, Malawi; Georgetown University, Washington D.C.; National AIDS Commission, Malawi; Blantyre City Council, Malawi; Blantyre CSO Network Chair, Malawi; Blantyre District Health Office, Malawi

**Keywords:** HIV prevention, Community-led monitoring, Organizational Network Analysis, Civil Society

## Abstract

**Background:** Civil Society Organizations (CSOs) play a critical role in HIV prevention and service delivery in Blantyre, Malawi. Yet district-based public health leaders often lack a nuanced understanding of CSO connections, service offerings, and linkages to health facilities; information needed for effective district-level coordination and facility level uptake of HIV prevention resources.

**Methods:** We conducted an organizational network analysis (ONA) of CSOs providing HIV prevention support in Blantyre to identify connections between CSOs, the strength of those connections, and the services these organizations offer. We sampled organizations in September 2021 and November 2023. We used Kumu Software to interactively display the relationships among CSOs. We assessed CSO connections to health facilities to gain a comprehensive understanding of linkages that impact how HIV prevention services are delivered.

**Results:** We had a sampling frame of 227 organizations with a final sample size of 154 organizations. Ninety-two (60%) organizations had at least one connection with another organization and 146 (95%) had at least one connection with a facility. There is a hub of connected organizations that work together to provide HIV prevention services in Blantyre. Of the 35 repeat organizations from both data collections, 12 increased their connections to other organizations and 14 increased their connections to other health facilities over two years.

**Conclusion:** Knowledge of the highly connected organizational hub allows the government and partners to leverage these organizations to support and enhance HIV prevention service delivery. Tracking network changes networks over time can enhance collaboration among organizations to maximize resources based on the greatest need.

## Background

Global efforts to expand access to Human Immunodeficiency Virus (HIV) treatment have increased life expectancy and reduced transmission [1]. Nevertheless, reservoirs of infection remain and will persist unless we deliver effective primary HIV prevention [2, 3]. For example, Malawi still experiences approximately 16,000 new HIV infections each year, and recent population-based surveys indicate that Malawi joins other countries in struggling to reach goals for reducing new infections [5]. Blantyre, Malawi has made tremendous gains to achieve the UNAIDS 95-95-95 goals with 87.0% of people living with HIV aware of their status, 84.7% on ART, 81% are virally suppressed [5]. While Blantyre has made notable strides to reduce HIV incidence and prevalence, both remain relatively high. In 2022, 2,000 individuals of all ages were infected with HIV in Blantyre [4]. For adults in Blantyre, 15-49 prevalence remains at 9% [4], for adolescent girls and young women, 1.7%, and for ages 15-24, 4%, respectively [5]. To further reduce incidence, the country continues to introduce and scale the latest HIV prevention interventions including the oral PrEP modalities and injectable PrEP options including, long-acting Cabotegravir and twice-yearly, Lenacapavir [6].

Even with these improvements in HIV prevention service delivery and modality options, current health systems experience challenges in supporting the delivery, uptake, and effective use of HIV prevention products, especially for key populations, hinder the deployment of novel prevention products that could reduce the risk of HIV infection. Poorly coordinated HIV prevention delivery systems—particularly those linked to community outreach and delivery—present a barrier to addressing persistent risk and to controlling HIV epidemics over the long term. There is no standardized practice for how civil society and the formal healthcare sector interact in Malawi, which can result in systemic challenges surrounding linkage to care and treatment and potential risk for loss to follow-up [7, 8]. In Malawi, there is a national Non-Governmental Organizational (NGO) board that registers and tracks all NGOs operating within the country. However, there is no single governmental entity or registration system tracking community organizations, the services they offer and who they work with. This can create systematic challenges for decision-making at the district and national level as well as service delivery needs at the patient and community level.

However, Civil Society Organizations (CSOs) have proven to be instrumental to expand coverage of HIV prevention services [9, 10, 11]. According to guidance from Blantyre HIV community stakeholders, the term “CSO” is an overarching term for all non-governmental organizations (NGOs), community-based organizations (CBOs), and faith-based organizations (FBOs). Civil society and community-based service delivery channels play a critical role in HIV prevention service delivery [12].

Understanding how community-based organizations and the formal health system relate to one another presents potential opportunities to address barriers and enhance service delivery, especially to key populations [13, 14]. For example, through strategic coordination, CSOs can better complement and extend existing services and strengthen the overall delivery of HIV prevention services [13, 14]. In some instances, a CSO may be more accessible and user-friendly than a formal health facility or clinic [13, 15] and therefore, can be strategically leveraged to engage clients and either provide the service needed, if appropriate (e.g., condoms or counseling) or refer them to the nearest health facility for services such as PrEP or TB treatment.

However, the network of organizations delivering services outside the formal health system in Malawi is complex and not fully understood, leading to missed opportunities for effective and efficient delivery of products and services at community level. Social network theory hinges on the idea that the structure and density of connections affect how the network functions as a whole [16]. Hence, the more connections organizations have results in greater collaboration and improved service delivery for communities [17]. With this, we conducted an organizational network analysis (ONA) as a means to understand the complex networks of civil society relating to HIV prevention. An ONA provided a way to visualize the various types of connections within the informal and formal healthcare sectors to tailor approaches based on context and particular populations [18]. Our objective was to identify and document strengths and weaknesses of the network of HIV prevention organizations in Blantyre to enhance district-level coordination and leveraging of existing organizational resources.

## Methods

### Overview and Objectives

We conducted both a cross-sectional survey and an organizational network analysis to map a network of CSOs providing HIV prevention services in Blantyre with support from the Bill and Melinda Gates Foundation through the Blantyre Prevention Strategy (BPS) project. BPS is a health systems-based approach to HIV prevention aimed at building capabilities needed at district level to target risk, generate demand, effectively deliver current and emerging HIV prevention products and interventions, and support effective and sustained use of prevention products by the end user. We conducted data collection among CSOs, which included FBOs, NGOs, CBOs, and “other” for those organizations that did not identify as one of those three types. In Blantyre District in September 2021 and November 2023 to identify gaps in HIV service offerings, to visualize how organizations are connected, and to illustrate changes in the organizational network over time. To focus on this change over time, analysis and results focus on the second phase of data collection as well as differences between the first and second phases of data collection for overlapping organizations only. Part of these landscape changes included identification of those organizations that closed, opened, or maintained service delivery over the two years. Additionally, we collected data on the CSOs at two time points to understand how the landscape of community networks changed over time as well as with the implementation of the Blantyre Prevention Strategy.

The CSO network analysis had three main objectives:

1. Provide a comprehensive landscape of HIV prevention services delivered by NGOs, CBOs, and FBOs in Blantyre;
2. Visualize the network of CSOs, including how they relate to the formal healthcare system and each other; and
3. Identify changes in the landscape of organizational networks over time and the ongoing activities of the Blantyre Prevention Strategy to improve future service delivery.

### Survey Design

For the first round of data collection in 2021, we developed an exhaustive list of CSOs operating in Blantyre by creating a sampling frame of organizations identified through eight government-operated CSO registration systems. Currently, Malawi does not have a single unified list of CSOs and as a result, there are multiple systems that often have duplicates of the same organizations. We collected the names of every organization listed in these registration systems and following the removal of duplicates across systems, we identified a total of 332 organizations that worked in HIV prevention in Blantyre. To be included in the sample, an organization had to be currently operating, providing HIV prevention services, and able to be contacted. We excluded 160 organizations that did not meet this inclusion criteria because they could not be traced, had closed their operation, could not be contacted, were not providing HIV prevention services, or refused to participate. For the second round of data collection in 2023, we built off the same list of 172 organizations from round 1, but also leveraged district-level stakeholder input to add emergent organizations to the list. This resulted in a list of 227 eligible organizations working in HIV prevention as a starting point for Round 2. Of the 227 organizations we sent a survey to, only 158 could be reached. After data cleaning, the final sample was 154. There were 35 overlapping organizations between the two rounds. Figure 1 demonstrates the flow of the final sample of organizations. Key organizations are highlighted in the results with organizational permission.

**Figure 1.**
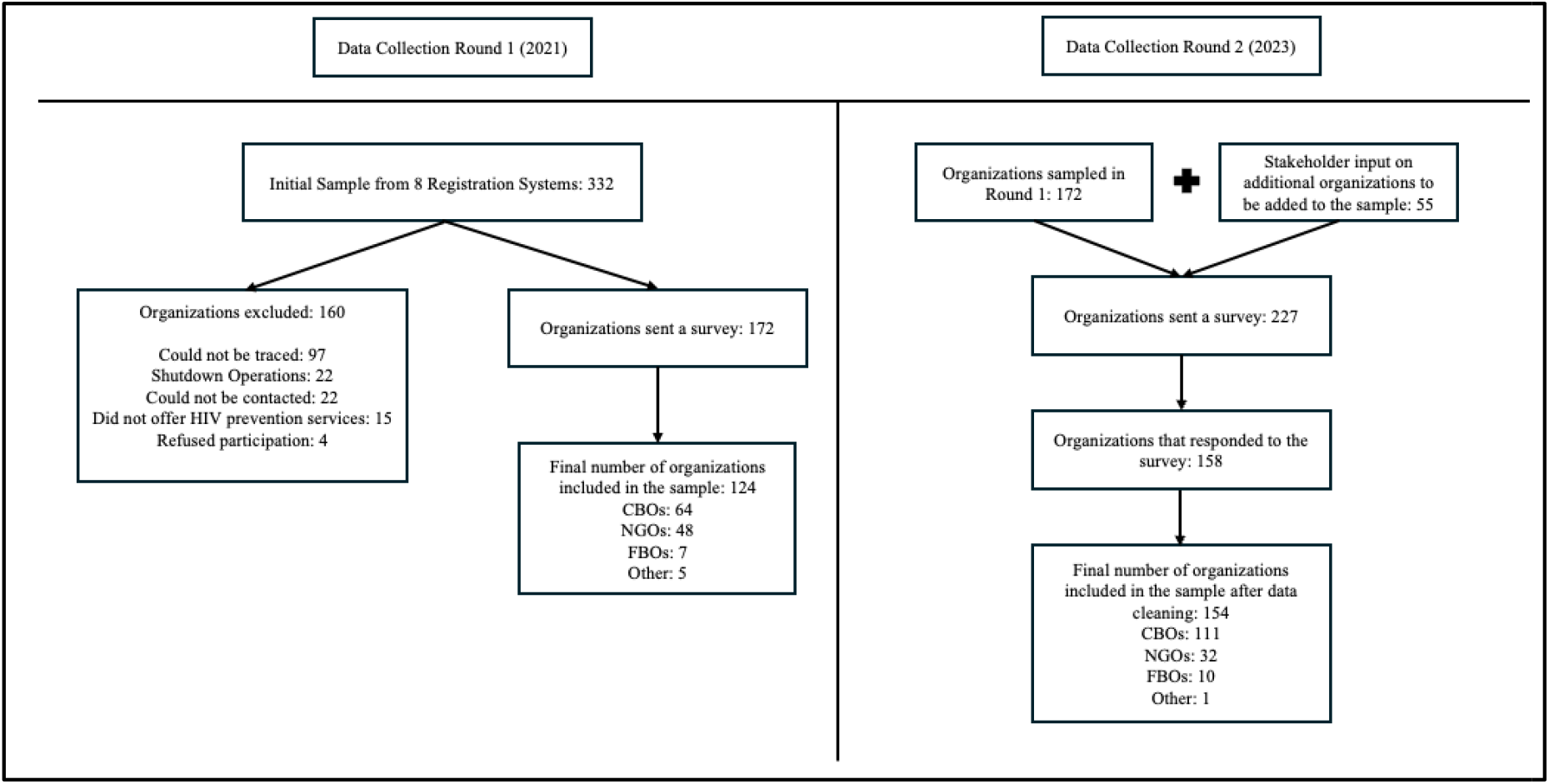
CSO Sample Collection Flow Chart

We categorized the survey into 5 sections:

1. <u>Administrative and funding data:</u> Questions focused on number of employees, office location, main sources of funding, and key donors.
2. <u>National Strategic Plan (NSP) and Local Authority HIV and AIDS Reporting Form indicators:</u> Questions focused on the NSP focus areas such as condoms and lubricant, pre-exposure prophylaxis (PrEP), key populations, elimination of mother-to-child transmission (eMTCT), sexually transmitted infections (STI), sexual and reproductive health (SRH), voluntary medical male circumcision (VMMC), and TB/HIV.
3. <u>Organizational network data:</u> Questions asked CSOs whether and what type of connection they had with other CSOs and health facilities. The types of connections explored between CSOs included sharing client referrals, information, commodities, and joint programming.
4. <u>HIV prevention services provided:</u> Questions asked which communities/populations benefited from services from among the general population and key populations, including men who have sex with men, female sex workers, and the Lesbian, Gay, Bisexual, Transgender, Queer, Intersex (LGBTQI) community. Questions on services were related to community mobilization and sensitization meetings, human-rights barriers, SRH / HIV prevention services, wellness & HIV prevention, condoms, STI screenings, PrEP, and TB testing and treatment.
5. <u>Open-ended questions asked about connections and organizational data collection and use.</u>

### Data Collection

For both rounds of data collection, we sent eligible organizations an online survey using Kobo Toolbox. <u>KoBoToolbox</u> is a free, open-source tool for creating surveys intended for data collection. While Kobo Toolbox does require data to submit the survey, data is also stored offline, so if respondents lose connection, their survey, will upload once connection is reestablished. All respondent organizations were offered data bundles to provide access to the online survey. Participants completed the survey using a phone, tablet, or computer. For the first round, organizations completed the surveys independently but could request technical support from our team of enumerators. Enumerators followed up with organizations that had not completed the survey within two weeks of receipt. For the second round, stakeholders from the city and district offices, including data clerks, the CBO network chair and others, were trained on the tool and supported the data collection. City and district staff both called and visited hard to reach organizations that had limited capacity to fill out the survey. Ultimately, this resulted in increased support for organizations to complete the survey as well as increased submissions by harder-to-reach organizations. We excluded surveys of organizations that could not be traced, had closed their operations, had unconfirmed contact information, or did not provide HIV services.

### Data Analysis

We used two approaches to analyze the survey data from CSOs:

1. Quantitative analyses using R Studio version 4.2.3, Microsoft Excel, and Tableau to generate descriptive statistics and visuals of the services being delivered by the CSOs.
2. An organizational network analysis (ONA) using Kumu. Kumu is an open-source online tool that helps display relationship data. We used the network data provided by respondents to display the relationships they have with other organizations as well as health facilities. The visuals created in Kumu also stratify connections based on organizations’ sharing client referrals, information, commodities, joint HIV programming, and the reliability of the relationship. We looked at the counts of connections and the frequency of organizations with no connections. Some organizations proved to be highly connected and central to the network, which resulted in an overall hub of organizations that were all connected. We note these highly connected organizations in the results with organizational permission to do so.

Network data assessed whether an organization worked with any other organization(s) and whether they worked with facilities. These data were used to understand the network of organizations offering HIV prevention services in Blantyre. The following metrics were assessed to understand the network:

- # of connections an organization had with other organizations,
- # of connections an organization had with public/private health facilities/centers,
- whether organizations shared clients and/or client referrals,
- whether organizations shared information,
- whether organizations shared commodities (sharing resources such as office space, written materials, pamphlets, posters, supplies, agricultural instruments, and/or equipment),
- whether organizations shared financial resources, and
- the nature of the relationship (e.g., strong, good, fair, poor relationship).

## Results

### Objective 1: Comprehensive landscape of HIV prevention services delivered

The following results focus on the second phase of data collection conducted in 2023. 154 unique organizations submitted a survey and were included in the analysis. The organizational breakdown is as follows: 111 CBOs (72%), 32 NGOs (21%), 10 FBOs (6%), and 1 other (1%). A quarter of organizations classify themselves as youth organizations where they offer services to children and adolescents and 95% of organizations indicated they work with a public or private health facility. Most organizations (105) have been operating for 10-plus years.

We also asked organizations about the number of staff and volunteers they employ and the amount of funding they receive on an annual basis (Table I). Most organizations (49) had a small annual budget of <$50 USD followed by a range of $50 to <$250 USD (35 organizations). Most organizations operate with a minimal budget and indicated “other” (40%) or international NGO (37%) as their main source of funding. In the “other” funding category, organizations indicated self-funding, donations, and fundraising as sources of funding. This aligns with organizational dependence on volunteers as the main type of staff.

In addition to funding and organizational structure, we focused on the types of HIV services organizations offered to clients (Table 1). Most CSOs provide messaging on HIV prevention (93%), condoms (78%), and referrals to health facilities for PrEP (34%). Conversely, very few CSOs indicated that they offer STI treatment (8%), VMMC delivery (7%), or PrEP (6%) (Table 1).

**Table 1.** Descriptive statistics of CSO staffing, budget, HIV prevention services offered, and key populations reached.

| Category | Sub-Category | CBO<br>(n=111) | FBO<br>(n=10) | NGO<br>(n=32) | Other<br>(n=1) | All<br>(n=154) |
| --- | --- | --- | --- | --- | --- | --- |
| Average # of Staff | Full-Time | 2 | 8 | 67 | 0 | 16 |
|  | Part-Time | 2 | 2 | 18 | 0 | 6 |
|  | Volunteers | 24 | 15 | 35 | 5 | 25 |
| Budget (USD) | <\$50 | 46 (30%) | 1 (1%) | 2 (1%) | 0 (0%) | 49 (32%) |
| | \$50 to <\$250 | 29 (19%) | 2 (1%) | 3 (2%) | 1 (1%) | 35 (23%) |
| | \$ 250 to <\$1,000 | 16 (10%) | 2 (1%) | 2 (1%) | 0 (0%) | 20 (13%) |
| | \$1,000 to <\$2,000 | 10 (6%) | 3 (2%) | 2 (1%) | 0 (0%) | 15 (10%) |
| | >\$2,000 | 10 (6%) | 2 (1%) | 23 (15%) | 0 (0%) | 35 (23%) |
| Services Offered | Messages to clients on HIV prevention | 104 (68%) | 10 (6%) | 28 (18%) | 1 (1%) | 143 (93%) |
|  | Condoms | 93 (60%) | 2 (1%) | 24 (16%) | 1 (1%) | 120 (78%) |
|  | Client referrals to health facilities delivering PrEP | 32 (21%) | 1 (1%) | 18 (12%) | 1 (1%) | 52 (34%) |
|  | SRH/HIV prevention services | 24 (16%) | 2 (1%) | 19 (11%) | 1 (1%) | 46 (30%) |
|  | TB services | 24 (16%) | 3 (2%) | 16 (10%) | 1 (1%) | 44 (29%) |
|  | Client referrals to health facilities delivering VMMC | 24 (16%) | 0 (0%) | 12 (8%) | 0 (0%) | 36 (23%) |
|  | HIV testing and counseling | 13 (8%) | 1 (1%) | 10 (6%) | 1 (1%) | 25 (16%) |
|  | eMTCT services | 11 (7%) | 1 (1%) | 8 (5%) | 1 (1%) | 21 (14%) |
|  | Workplace wellness and HIV prevention programs | 6 (4%) | 1 (1%) | 6 (4%) | 1 (1%) | 14 (9%) |
|  | STI treatment | 3 (2%) | 1 (1%) | 7 (5%) | 1 (1%) | 12 (8%) |
|  | Lubricant | 3 (2%) | 0 (0%) | 8 (5%) | 0 (0%) | 11 (7%) |
|  | PrEP Dissemination | 0 (0%) | 0 (0%) | 9 (6%) | 1 (1%) | 10 (6%) |
|  | VMMC Delivery | 7 (5%) | 1 (1%) | 3 (2%) | 0 (0%) | 11 (7%) |
| Key Populations | FSWs | 35 (23%) | 1 (1%) | 18 (12%) | 0 (0%) | 54 (35%) |
|  | MSM | 3 (2%) | 0 (0%) | 5 (3%) | 0 (0%) | 8 (5%) |
|  | LGBTQI | 3 (2%) | 0 (0%) | 3 (2%) | 0 (0%) | 6 (4%) |

We found that only 56 (36%) CSOs offered HIV prevention services to key populations. The number of CSOs providing services for key populations is as follows: 54 (35%) organizations offer to female sex workers (FSWs), 8 (5%) organizations offer to men who have sex with men (MSM), and 6 (4%) organizations offer to LGBTQI. The specific services offered to each population can be found in Figure 2. STI screening and testing are the least offered services to all key populations indicating a potential gap in services for STIs.

**Figure 2.**
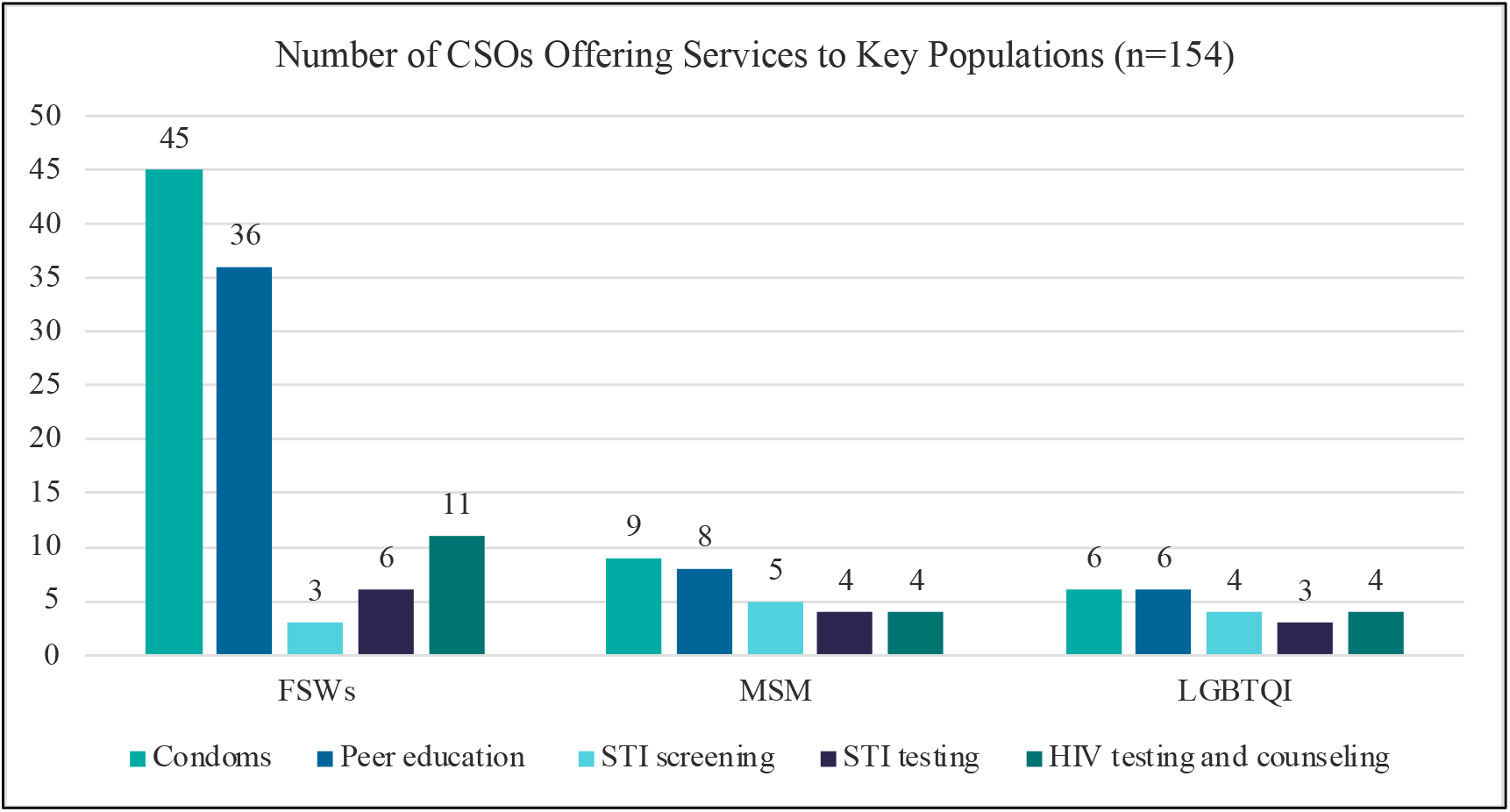
HIV prevention services offered to key populations

### Objective 2: Visualize the network of CSOs with one another and to the formal healthcare network

The network maps in Figure 3 display connections between CSOs and health facilities with the nodes not sized by any characteristics. A cluster of organizations emerged, shown in the center of the map (Figure 3. Connections between CSOs), which demonstrates the existence of a highly connected network of organizations working together in Blantyre. Ninety-two (60%) organizations indicated that they had at least one connection with another organization. Despite this cluster of connected organizations, 62 (40%) CSOs do not have a relationship with another CSO and operate independently (Figure 3. Connections between CSOs).

**Figure 3.**
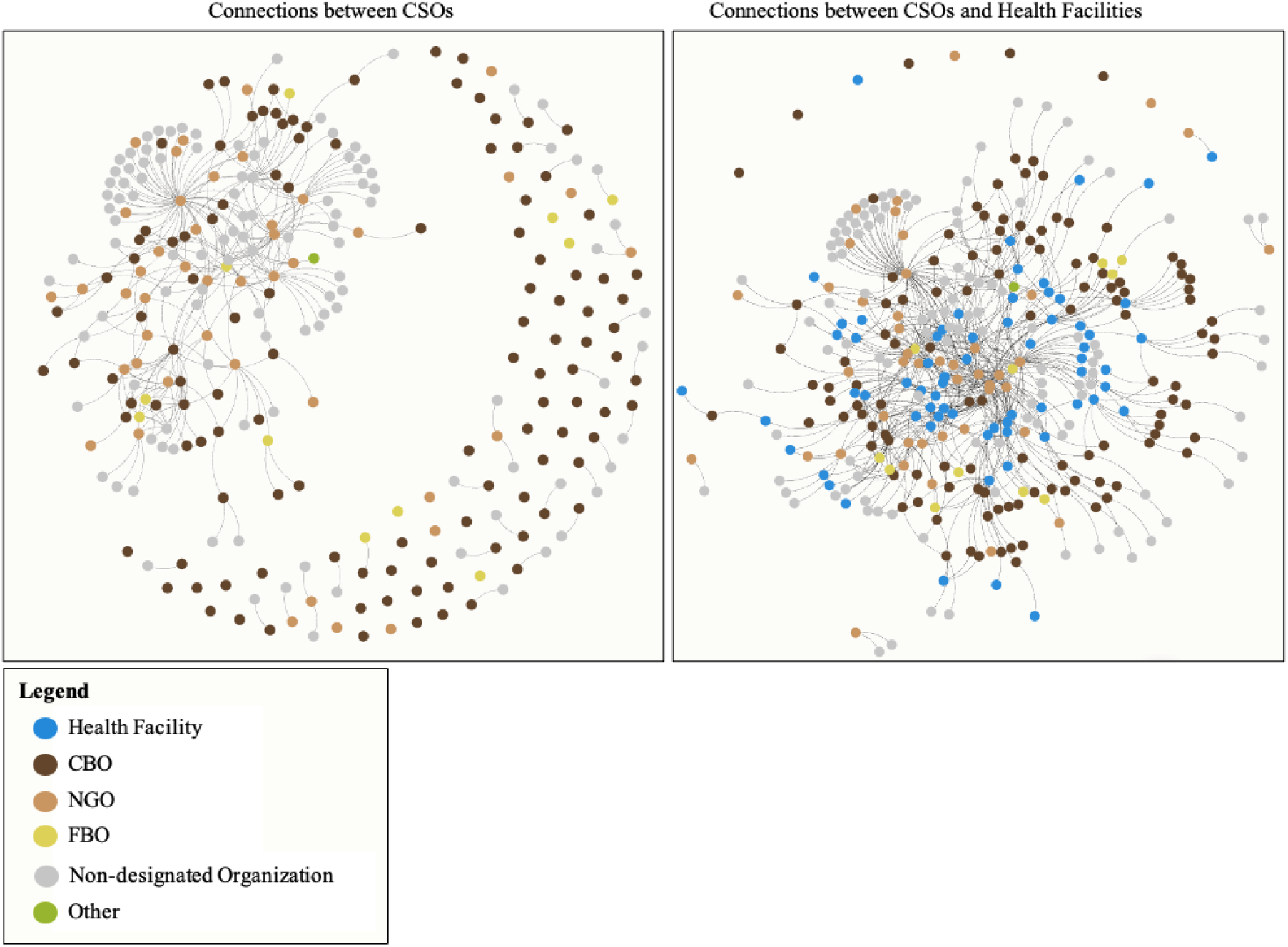
Network Maps between CSOs and Health Facilities

Though CSO to CSO connections are just over 50%, CSOs are highly connected to health facilities. 95% of organizations indicate a connection to a health facility (Figure 3. Connections between CSOs and Health Facilities). CSOs are also more likely to have more than one health facility partner rather than multiple organizational partners. Only 24% of CSOs reported more than 1 CSO connection, whereas 38% of organizations reported more than 1 facility partner. Only 8 (5%) organizations said they did not work with a health facility in any capacity.

The following key terminology for network analyses is helpful to interpret the outputs and findings of the CSO ONA mapping. These include:

- Cluster: a set of densely connected nodes.
- Connection: a line between two nodes indicating a relationship between a CSO and another organization.
- Network: the organizations that make up the mapping (i.e., respondent organizations and another organization).
- Node: a circle that represents a CSO, health facility, or another organization.
- Organizational Network Mapping: a way to summarize and visualize the relationships between organizations.
- Periphery/Outskirts: refers to the outside area of the map where nodes with very few or no connections are shown. This shows organizations that are siloed and not well-connected.

#### Types of Connections and Relationship Quality

For those organizations that indicated they were connected to other CSOs, joint HIV programming (56%) was the most common type of connection between organizations. Joint HIV programming was when organizations conduct activities and programs related to HIV prevention or awareness together. Information sharing (50%) (sharing of ideas or thoughts on resourcing/clients) was the second most common type of connection, followed by commodity sharing (40%) (sharing resources such as office space, written materials, pamphlets, posters, supplies, agricultural instruments, health commodities, and/or equipment). Client referrals were not a common form of organizational connection (35%); however, financial sharing, defined as the giving or receiving of funds, was the most uncommon form of connection between CSOs (4%) (Figure 4).

**Figure 4.**
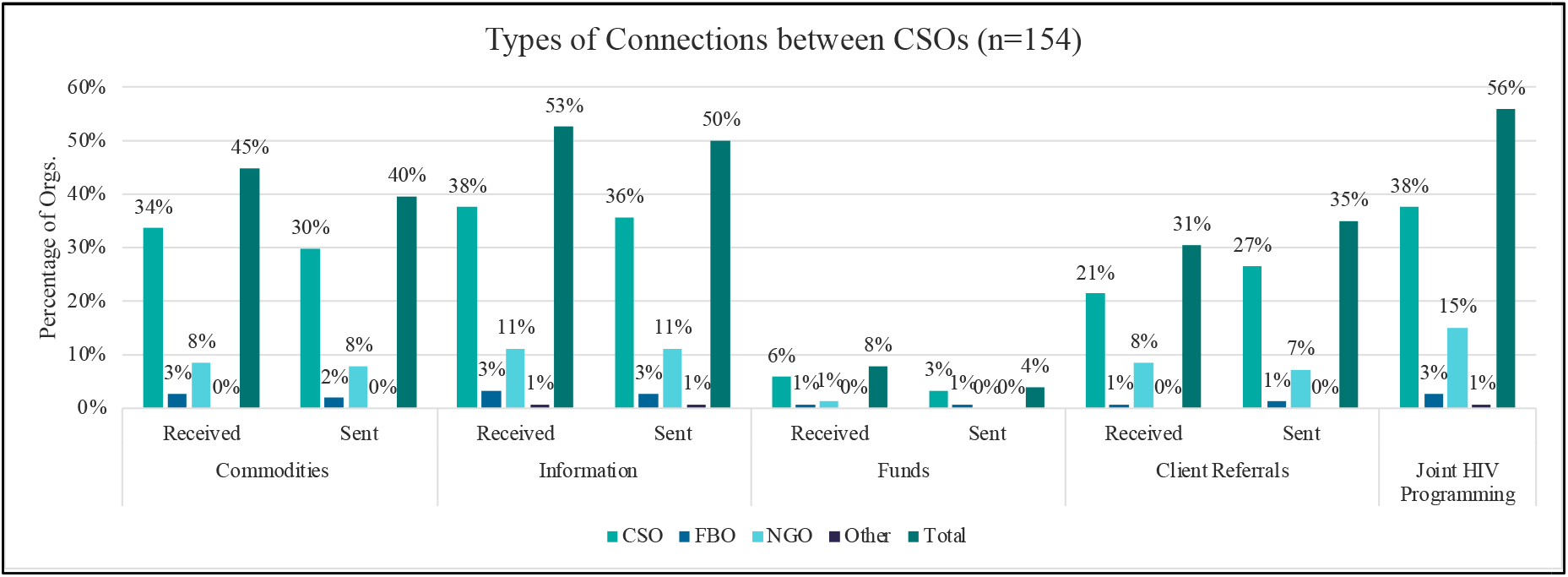
Frequency and Type of Connection between CSOs

### Joint HIV Programming, Commodity Sharing, and Information Sharing

Organizations indicated that they often conduct joint awareness campaigns or mobilization events with other organizations in their communities. 38% of CSOs and 15% of NGOs indicated they conduct joint HIV programming with other organizations (Figure 4).

Information sharing was the second most common type of connection between organizations. Fifty-three percent of respondents have a partner they received information from. Similarly, organizations sent information to 50% of their connected CSOs. CSOs shared information with other organizations with the following frequency: never (15%), less than once a month (23%), 1-3 times per month (26%), 1-2 times a week (1%) more than two times per week (3%). Figure 5 below demonstrates those CSOs who share information with partner organizations. The nodes are sized by the number of connections: the larger the node, the more connections an organization has. Organizations are highly connected by way of information sharing.

**Figure 5.**
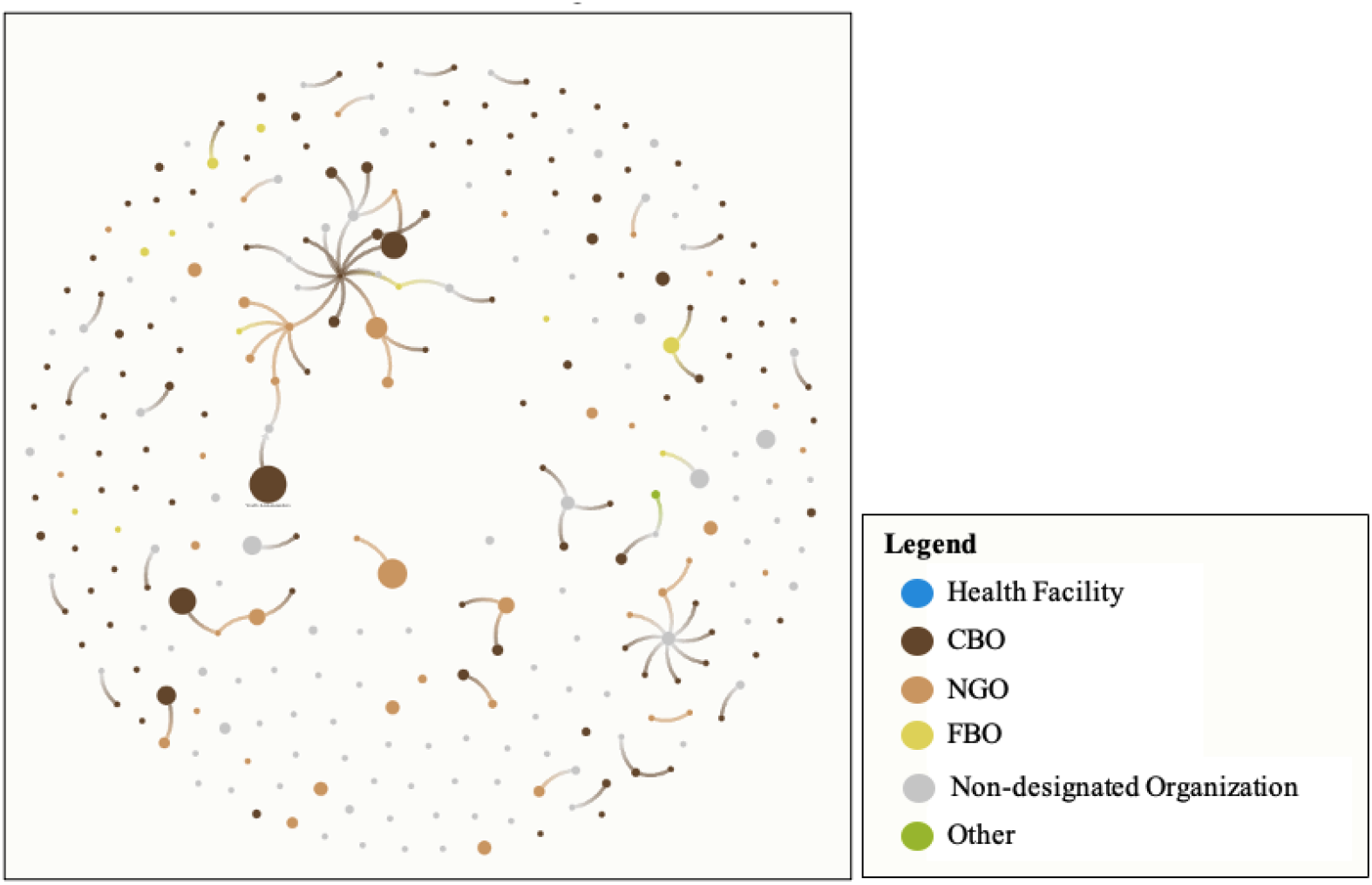
CSOs send information to another organization

Note: node size is determined by the number of connections of an organization. The largest node has the greatest number of connections (e.g., 12 connections). The smallest node has zero connections.

Similarly, 45% of CSOs indicated they received commodities of some sort from other organizations and 40% said they sent commodities to other organizations.

### Financial Sharing and Client Referrals

Conversely, client referrals and financial sharing were the most uncommon forms of relationship sharing between organizations. Organizations shared very little financial resources with other organizations. Only six organizations (4%) shared funds with another organization, the amount was <$1,000 USD. Given that most organizations are funded by other international organizations/donors, it is not surprising that this category is the lowest type of sharing. Referrals sent and received between CSOs were also rare. Thirty-seven percent of CSOs never received client referrals from another CSO and 32 percent never sent referrals to another CSO. This shows that CSOs are even less likely to receive referrals from their connections than send referrals to their connections. When referrals do take place, it is almost always less than 100 referrals per year. Client referrals are shown in Figure 6 below. Connections are sized by the number of referrals - a thicker line indicates more referrals sent.

**Figure 6.**
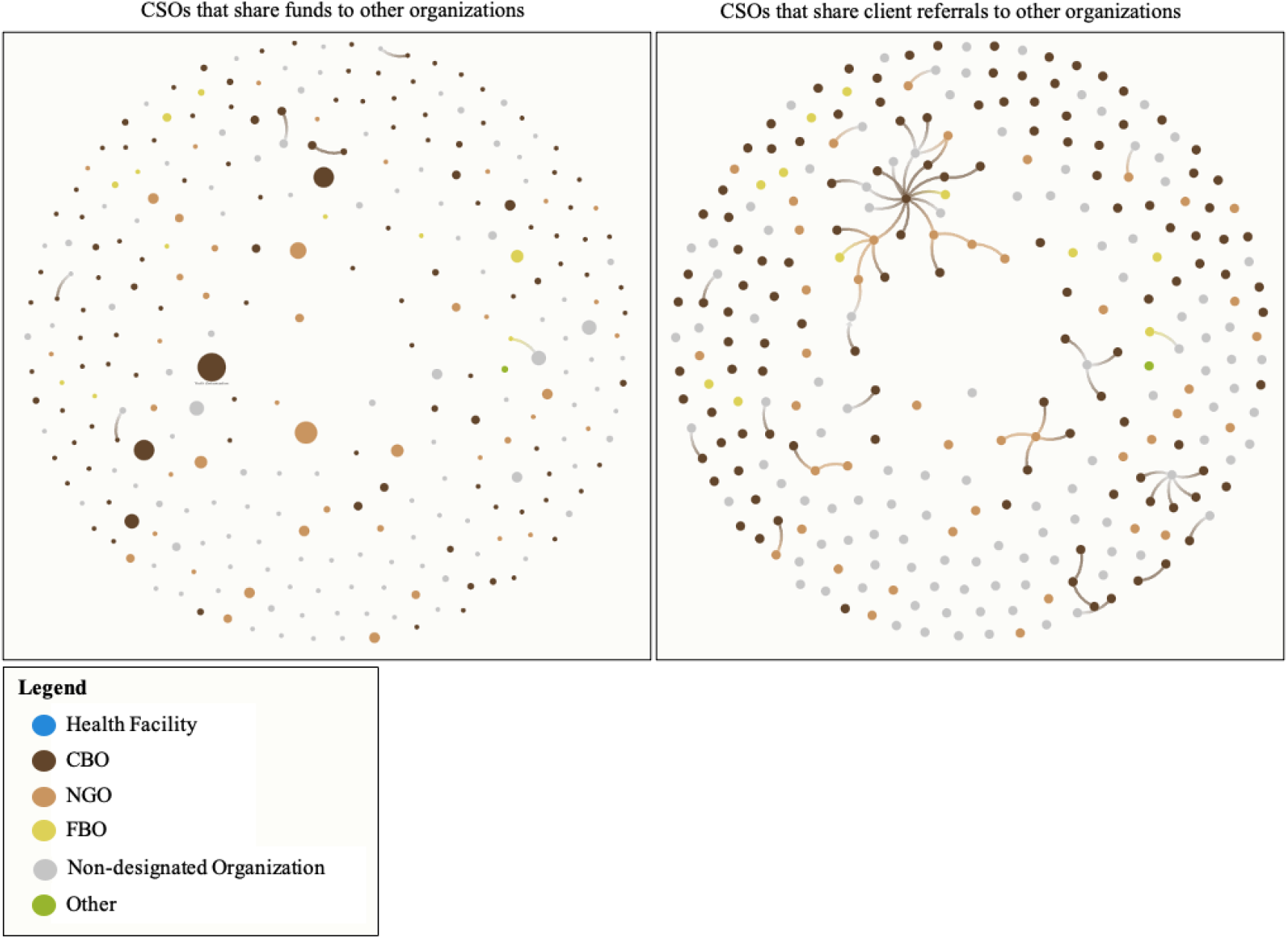
CSOs that share funds (left) and send client referrals to other organizations (right)

### Relationship Quality

In Figure 7, a cluster or set of densely connected nodes emerges when organizational connections are displayed by relationship quality. Within the cluster, 37% of organizations reported an ‘excellent’ (10%) or ‘good relationship’ (28%) between organizations, while 30% reported ‘fair’ (26%) or poor (5%) relationships. Of those 38% of organizations that indicated they had an excellent or good relationship with connected organizations, all but 8 (14%) indicated they do joint HIV programming with other organizations. Of those organizations that indicated they had a poor relationship, or those characterized by little reliability with connected organizations, none sent funds, most did not do any client referrals or minimal clients referrals (<100 clients), and did not share information frequently (either never or <3x per month).

**Figure 7.**
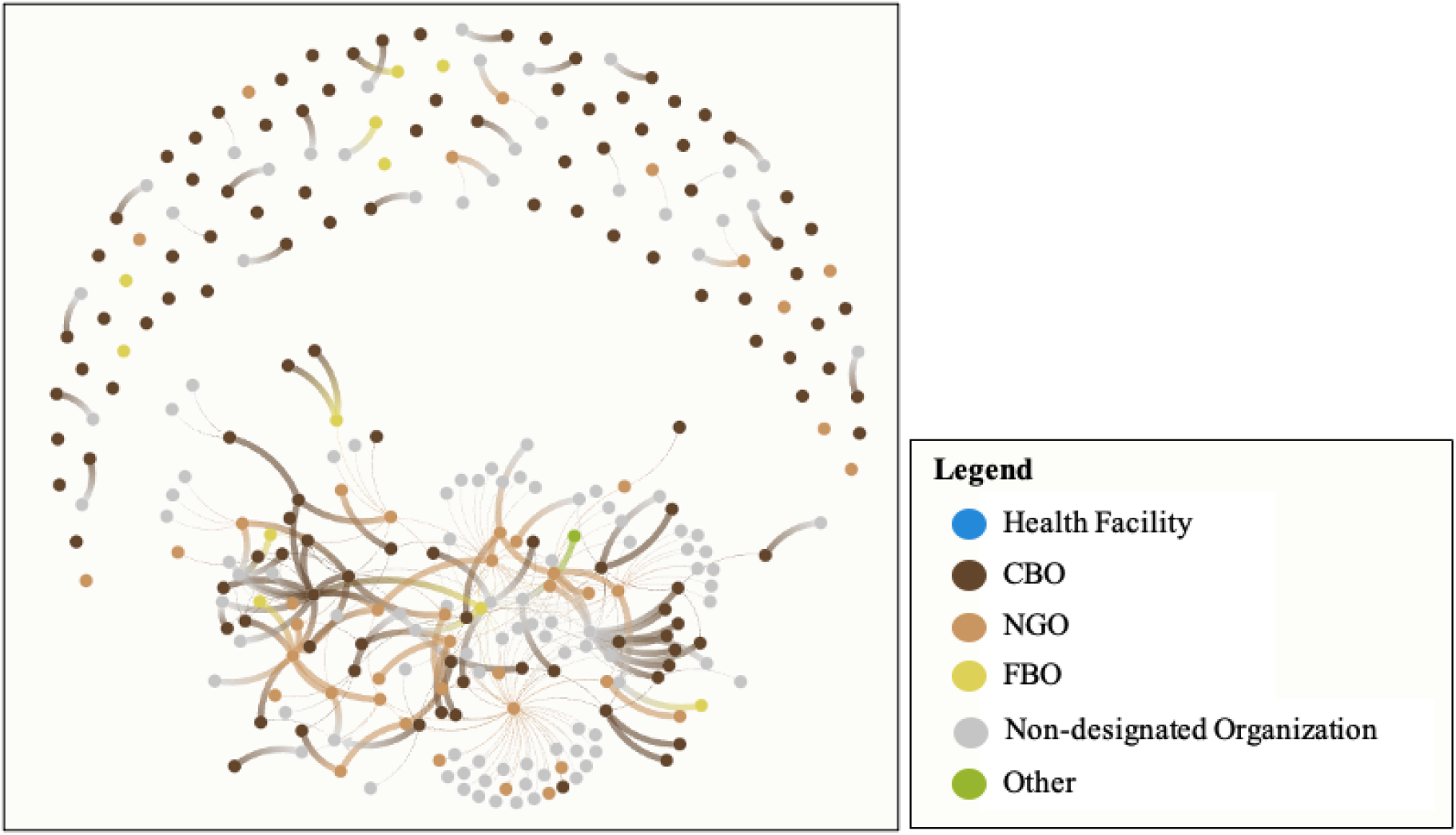
The quality of relationships between CSOs shown by line width (thicker line = highly reliable relationship)

### Objective 3: Identify changes in the landscape of organizational networks over time and the ongoing activities of the Blantyre Prevention Strategy to improve future service delivery

Only 35 organizations participated in both rounds of data collection, due to the availability and response of organizations and changes to the organizational landscape over time. Given the limited number of overlapping organizations between the two rounds, we acknowledge the limitations in conclusions that can be drawn over time. Comparing the 35 organizations helped us to understand changes to the landscape. The emergence of many new organizations demonstrates heterogeneity in the landscape. However, of those that responded two both surveys, we identified changes in their operations over time.

Overall, 14 of 35 organizations reported an increased number of connections to health facilities and 12 organizations increased the number of connections to CSOs in the 2nd data collection (2023) compared to the first (2021) (Table 2). Six CSOs reported an increase of 1 connection, five CSOs reported an increase of 2-5 connections, and three organizations reported an increase of more than 5 connections to health facilities. Three CSOs reported an increase of 1 connection, five CSOs reported an increase of 2-5 connections, and four CSOs reported an increase of more than 5 connections to other organizations in phase 2.

**Table 2.** The change in number of CSO connections to health facilities and other CSOs from the 1^st^ to the 2^nd^ data collection.

| 1<sup>st</sup> to 2<sup>nd</sup> data collection (n = 35) | Health Facilities | CSOs |
| --- | --- | --- |
| Increase | 14 | 12 |
| Decrease | 9 | 13 |
| Static | 12 | 10 |

Nine organizations reported decreased linkages to health facilities and 13 organizations saw a decrease in the number of connections to other CSOs in the 2nd collection (Table 2). Finally, 12 organizations reported the same number of connections to health facilities and 10 organizations reported the same number of connections to CSOs (Table 2).

Figure 8 displays CSO hubs or those organizations with a high number of connections and their changes from the 1st CSO mapping to the 2nd CSO mapping. Chitzanso Youth Support Organization (CHIYOSO) proved to be one of the most connected organizations and central to the CSO network. In the 1st data collection, CHIYOSO was connected to 14 other CSOs and 6 health facilities (Figure 8 top left). In the 2nd data collection, CHIYOSO doubled its number of connections to health facilities (12) and slightly decreased its connections to other CSOs (12) (Figure 8 top right). In Figure 8, node location is not relevant, only the line connection and node size are pertinent. In the 1st round of data collection, Youth Ambassadors was the second most connected organization with connections to 7 health facilities and 12 other CSOs (Figure 8 bottom left). Youth Ambassadors recorded a decline in overall connections in the 2nd round of data collection with 4 health facilities and 5 CSOs (Figure 8 bottom right).

**Figure 8.**
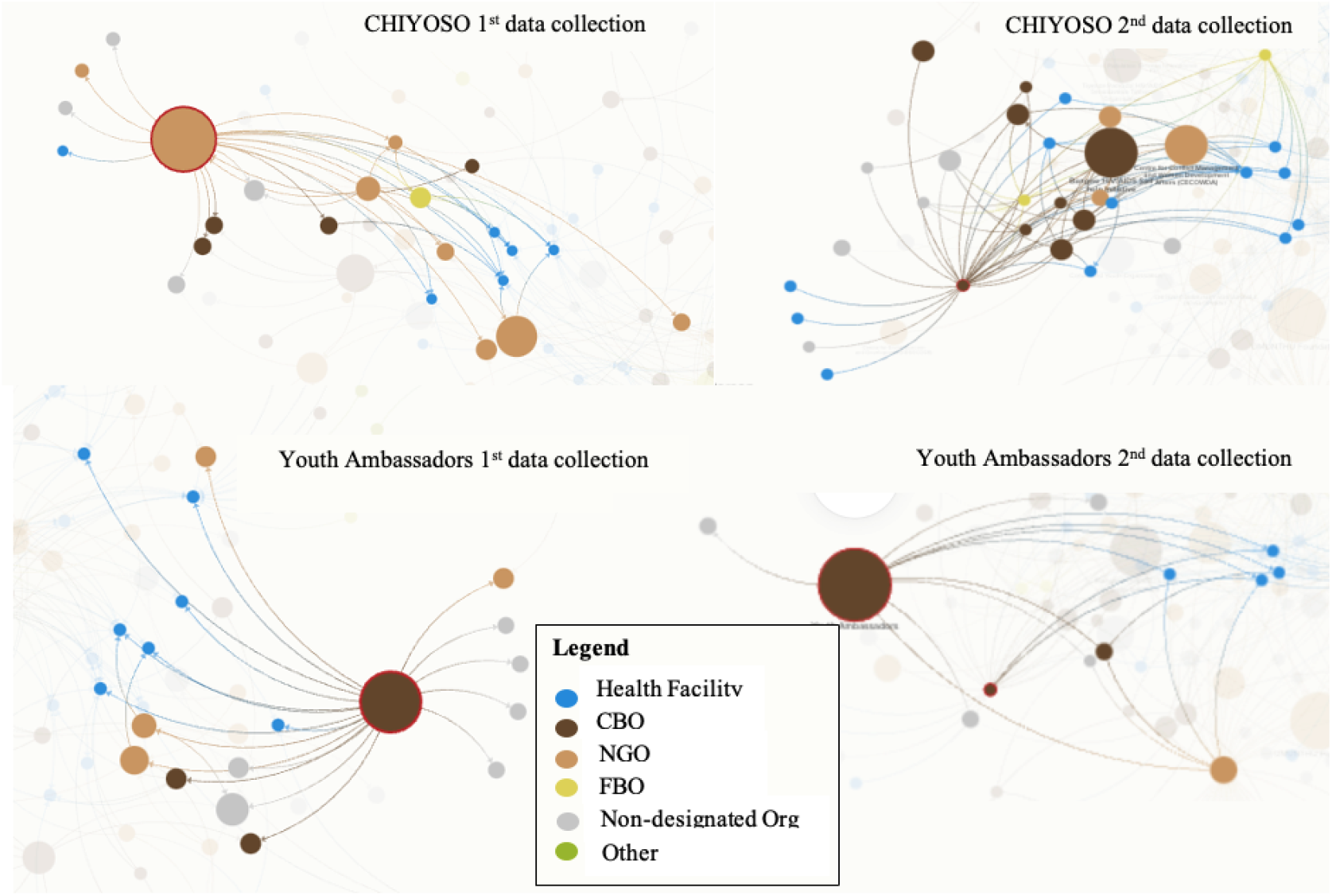
CSO hub examples in the 1st data collection vs. 2nd data collection

## Discussion

The organizational network analysis illuminated a central network of community organizations offering community-based HIV prevention services in Blantyre, but also identified lack of coordination between organizations as so many operate independently. Visualization of the connections among organizations enhanced programmatic awareness of services offered by CSOs and populations served (or lack thereof). We found a limited number of community organizations offering services to key populations like MSM and LGBTQI individuals. Additionally, we identified key organizations as hubs/highly connected, which is essential knowledge for District Councilors and City Councilors as they serve their respective constituents in Blantyre. Highly connected organizations were those shown in figure 8 that list many organizations they are connected to in some way. Knowledge of these hubs supports service delivery for high-risk clients. Additionally, well-connected organizations can serve as centers of information and resources for less-connected organizations through potential partnerships or community mobilization events.

This study highlights clear service gaps (lack of services offered) by community organizations such as limited STI (8%) and tuberculosis (TB) (29%) services as well as PrEP (6%) offerings (Table 1). Service gaps by CSOs for MSM and LGBTQI key populations were also high – only 5% of organizations offered services for MSM and 4% for LGBTQI (Table 1). This is where CSOs can leverage their connections to health facilities even if they are not very connected to other organizations. Knowledge of both the informal and formal networks, enhance overall patient care and coverage.

A well-connected CSO may not offer TB testing but may refer a client to another CSO or facility for a test. However, organizations that operate independently of the formal sector and other organizations may not have this information and client care may suffer as a result. Well-connected organizations often had more resources (e.g., higher operating budgets, more staff, more volunteers etc.) compared to siloed organizations. A well-connected organization could become a middleman by connecting organizations providing similar services together to avoid duplication of services. In Blantyre, one organization had a surplus of condoms but indicated they did not know what to do with the surplus or where to send them. This is where stakeholders in Blantyre can leverage the results of the network analysis through increased coordination of organizations, goods and services in the district.

The U.S. President’s Emergency Plan for AIDS Relief (PEPFAR) highlights that community-led responses and monitoring filtered through civil society provide communities a voice and mechanism for decision-making and allow other sectors to pinpoint remaining gaps and challenges to HIV service delivery, uptake, and retention [10, 11]. These analyses demonstrate the importance community organizations play in HIV service delivery, programming, and interventions. It is evident that CSOs can work with health facilities as well as provide an additional avenue to obtain the services they need for adequate HIV prevention and care both at other CSOs and at health facilities [19-21]. Awareness of how community-based organizations operate and work together in a communal network has important implications for creating a comprehensive care network for people living with HIV [22-23]. Further, if organizations also understand the network of HIV prevention organizations they operate within, they can have increased awareness of the gaps in HIV prevention services, and potentially fill that gap rather than duplicate services. Competition for funding and resources is a key aspect to consider amongst organizations, but knowledge of the gaps provides an opportunity for additional sources of funding as opposed to fighting for the same resources for the same services.

The ONA provides a systematic way to understand a very heterogeneous and amorphous section of the health care system within a country that has the potential to increase awareness of support systems, increase access to treatment and care, and reduce the overall disease prevalence in an area [24, 25, 26]. An ONA is less commonly used in a community-based context and thus, is a unique methodological tactic that can be leveraged as a tool by community leaders and members to strengthen relationships and leverage resources between the informal and formal healthcare sectors [25, 25]. Routine data on the organizational network can indicate critical opportunities and gaps for innovative program design, influence resource allocation, and monitor strategy pivots [24]. Capitalizing on those organizations at the center of networks leverages civil society to more effectively reach target populations with more comprehensive prevention services [24, 27-28]. Conversely, working with CSOs operating in silos can increase collaboration, decrease duplication, and identify organizations well-positioned to deliver novel HIV prevention technologies as they are deployed.

### Recommendations

Well-connected CSOs play an integral role in the community for clients to access comprehensive HIV prevention services. The following recommendations emerged from this analysis to enhance the coordination of organizations and improve HIV prevention service delivery in Blantyre. 1) Leverage the reach of CSOs in the community to provide messaging and information on sensitization, outreach, and mobilization, especially for key and vulnerable populations. 2) CSOs should capitalize on their connections to health facilities. Our findings demonstrate that most CSOs lack certain service offerings like demand generation for PrEP, STI screening/treatment and TB testing/treatment. Connections with health facilities can fill the gap in service delivery that CSOs do not offer. 3)Strengthen coordination and connections between CSOs to maximize resources and service delivery across the HIV prevention network. Strengthened connections between CSOs could help ensure service delivery is complementary and individuals can access a comprehensive set of services. 4) Interventions that aim to increase CSO HIV prevention service delivery and strengthen CSO connections should focus on increasing referrals between CSOs, especially those that offer complementary services. CSOs typically specialize in specific service areas as it is costly to offer a comprehensive set of services. This specialization makes CSO connections crucial; collaborations among these organizations spur the synergy that is integral to strengthening service delivery. Referrals between CSOs are especially important for ensuring individuals can access the HIV prevention services they need within the community. This analysis demonstrates areas of services like STI treatment and VMMC delivery that require clients to seek at facilities or other organizations, which CSOs can support clients to access. 5) Develop a unified national-level registration system that houses all organizations.

### Sustainability

While this tool was successfully deployed in Blantyre, Malawi, it could also be applied to other contexts to display the relationships of the civil society network as well as provide a landscape analysis of the formal and informal sectors. An ONA can be conducted at local (e.g. village/community) and central/national levels(nationally) in Malawi as well as other countries as exemplified by its use in both Addis Ababa, Ethiopia and Chicago, US [18, 29].

The ONA can be institutionalized at various levels of analysis and used routinely to assess how the web of support shifts over time and can be further optimized to enhance care and service delivery [24]. This type of mapping can be used routinely to monitor changes over time in established connections as well as new additions to the network to fully understand and capture service delivery offerings in a community. This helps identify gaps in services to date and can guide program planning and implementation of services and technologies [24-26]. Program success and sustainability are only possible when connections are understood within a community [28, 21]. An ONA can be applied in a similar way to monitor program success over time [28].

Key actors in Blantyre see the CSO network as playing an integral role in the scale-up of PrEP and other HIV prevention technologies, given how embedded CSOs are in the community. CSOs can provide an additional service delivery network outside of the formal sector for HIV prevention services through the community, increasing demand generation, and enhancing sensitization around these services.

### Limitations

Some CSOs may not have been captured if they were not listed in the 8 government registration systems used to develop the list of organizations. This study could be enhanced with additional qualitative analyses. For example, further insights would be helpful to understand how and why an organization characterizes a relationship as good or excellent. Further, during the sampling phase, issues arose in tracking down organizations listed in the registers - either due to outdated registers or organizations closing or not operating, reducing the overall sample size. The small sample of 35 overlapping organizations between round 1 and round 2 limited the types of comparisons we could make overtime. The low number of overlapping organizations was due to several factors including some organizations shutting down, changed contact information, and refusal to participate. This also highlights some challenges in tracking service delivery in the informal healthcare sector in LMICs.

### Conclusion

This analysis highlights strong heterogeneity among CSOs supporting HIV prevention in Blantyre. Conducting the ONA regularly could help improve connections between the formal and informal healthcare sectors by being able to identify those CSOs operating independently and linking the other organizations and facilities. Additional ONAs can also support program planning for HIV prevention services provided, understand CSO catchment areas and reach, and identify ongoing gaps in service coverage. Involving key stakeholders from the city and district levels in Blantyre was key to ensuring this process was a locally led approach. With Blantyre District and Blantyre City key stakeholders actively involved in the second data collection, this could promote continued data collection on a yearly or biannual basis to obtain the most recent data on community network changes. The results from the network analyses have been used by stakeholders and the National AIDS Commission to inform their work of which organizations are operating and which organizations in specific areas can service identified. Having up-to-date, well-documented, and pre-mapped systems can help increase and maintain access to services for priority populations and be one solution to addressing both the ethical challenges of new HIV prevention technologies as well as dealing with environmental and social hazards of seeking care. Understanding the landscape of HIV prevention community networks by key stakeholders provides an opportunity for those with decision-making power to ensure people living with HIV can access care and treatment both at community organizations and at health facilities.

## Data Availability

All data produced in the present study are available upon reasonable request to the authors.

## Declarations

## Ethics approval and consent to participate

All ethical clearance was approved and obtained by the National Health Research Ethics Committee in Malawi. Additionally, all organizations provided digital consent before filling out the questionnaire.

## Consent for publication

Not applicable as all data are the organizational level.

## Availability of data and materials

The datasets generated and analyzed during the current study are not publicly available due to limitations of data sharing agreements under the overarching project but are available from the corresponding author upon reasonable request.

## Competing interests

The authors declare that they have no competing interests.

## Funding

This work was funded by the Gates Foundation and implemented by Georgetown University.

## Authors’ contributions

RH is the corresponding author and led survey design, data management, data analysis, and manuscript writing. PK supported the design of the work as well as data collection and manuscript review. YK supported the design of the work, district coordination to collect data, and manuscript review. CM supported data collection and manuscript review. RN supported data collection and manuscript review. SM supported data collection and manuscript review. DG supported data analysis and manuscript review. TS supported the design of the work and manuscript review. HC supported the design of the work as well as data collection and manuscript review. CH supported the design of the work as well as data collection and manuscript review. SA supported the design of the work as well as data collection and manuscript review. DH supported the design of the work as well as data collection and manuscript review. GK supported the design of the work as well district coordination and collaboration.

## Acknowledgements

Not applicable.

## List of Acronyms

ART: Antiretroviral Therapy
CBO: Community-Based Organization
CHIYOSO: Chitzanso Youth Support Organization
CSO: Civil Society Organization
eMTCT: Elimination of Mother-to-Child Transmission
FBO: Faith-Based Organization
HIV: Human Immunodeficiency Virus
LGBTQI: Lesbian, Gay, Bisexual, Transgender, Queer/Questioning, Intersex
LMICs: Low- and Middle-Income Countries
MSM: Men who have Sex with Men
NGO: Non-Governmental Organization
NSP: National Strategic Plan
ONA: Organizational Network Analysis
PEPFAR: The U.S. President’s Emergency Plan for AIDS Relief
PLHIV: People Living with HIV
PrEP: Pre-Exposure Prophylaxis
SRH: Sexual and Reproductive Health
STI: Sexually Transmitted Infection
TB: Tuberculosis
UNAIDS: Joint United Nations Programme on HIV/AIDS
USD: United States Dollar
VMMC: Voluntary Medical Male Circumcision

## Notes

### Competing Interest Statement

The authors have declared no competing interest.

### Funding Statement

This study was funded by The Gates Foundation.

## References

1. PEPFAR. Results and impact - PEPFAR [Internet]. 2022 [cited 2023 Aug 10]. Available from: https://www.state.gov/results-and-impact-pepfar/

2. Akullian A, Bershteyn A, Jewell B, Camlin CS. The missing 27%. AIDS. 2017 Nov 13;31(17):2427–9. doi: 10.1097/QAD.0000000000001638. Available from: https://www.ncbi.nlm.nih.gov/pmc/articles/PMC5673304/

3. UNAIDS. The youth bulge and HIV [Internet]. Geneva: UNAIDS; 2018 [cited 2023 Aug 10]. Available from: https://www.unaids.org/en/resources/documents/2018/the-youth-bulge-and-hiv

4. PALMS. NAOMI Model 2023 predictions [Internet]. 2023 [cited 2023 Aug 2]. Available from: https://palms-staging.coopersmith.org/ext/index

5. Malawi Population-Based HIV Impact Assessment. Summary sheet [Internet]. 2020 [cited 2023 Jul 18]. Available from: https://phia.icap.columbia.edu/wp-content/uploads/2022/03/110322_MPHIA_Summary-sheet-English.pdf

6. Philbin MM, Perez-Brumer A. Promise, perils and cautious optimism: the next frontier in long-acting modalities for the treatment and prevention of HIV. Curr Opin HIV AIDS. 2022 Mar;17(2):72–88. doi: 10.1097/COH.0000000000000723.

7. Uberoi D, Ojo T, Sriharan A, et al. What cn implementation scieance offer civil society in their efforts to drive rights-based health reform?. Glob Health Res Policy. 2023;8:1–8. doi: 10.1186/s41256-023-00284-4.

8. McDonough A, Rodríguez DC. How donors support civil society as government accountability advocates: a review of strategies and implications for transition of donor funding in global health. Global Health. 2020;16:1–14. doi: 10.1186/s12992-020-00628-6.

9. Aaraj E, Chrouch MJA. Drug policy and harm reduction in the Middle East and North Africa: the role of civil society. Int J Drug Policy. 2016;31:168–71. Available from: http://www.globalcommissionondrugs.org/wp-content/uploads/2016/06/Special-Issue-MENA-GCDP-IJDP-1.pdf

10. UNAIDS. Final report on community-led AIDS responses based on the recommendations of the Multistakeholder Task Team [Internet]. 2022 [cited 2023 Aug 2]. Available from: https://www.unaids.org/en/resources/documents/2022/MTT-community-led-responses

11. PEPFAR. Community-led monitoring fact sheet [Internet]. 2020 [cited 2023 Jul 17]. Available from: https://www.state.gov/wp-content/uploads/2020/07/PEPFAR_Community-Led-Monitoring_Fact-Sheet_2020.pdf

12. UNAIDS. Supporting community-based responses to AIDS: a guidance tool for including community systems strengthening in Global Fund proposals [Internet]. Geneva: UNAIDS; 2009 [cited 2023 Aug 10]. Available from: https://www.unaids.org/sites/default/files/media_asset/20090218_jc1667_css_guidance_tool_en_0.pdf

13. World Health Organization. Health policy and system support to optimize community health work programmes for HIV, TB and malaria services: an evidence guide [Internet]. 2020 [cited 2022 Jan 10]. Available from: https://apps.who.int/iris/bitstream/handle/10665/340078/9789240018082-eng.pdf?sequence=1

14. Montenegro H, Holder R, Ramagem C, Urrutia S, Fabrega R, Tasca R, et al. Combating health care fragmentation through integrated health service delivery networks in the Americas: lessons learned. J Integr Care. 2011;19(5):5–16. doi: 10.1108/14769011111176707.

15. Auschra D. Barriers to the integration of care in inter-organisational settings: a literature review. Int J Integr Care. 2018;18:1–12. doi: 10.5334/ijic.3068.

16. Provan KG, Beagles JE, Leischow SJ. Network formation, governance, and evolution in public health: the North American Quitline Consortium case. Health Care Manage Rev. 2011;36(4):315–26.

17. Hjern B, Porter DO. Implementation structures: a new unit of administrative analysis. Organ Stud. 1981;2(3):211–27.

18. Thomas JC, Reynolds H, Bevc C, Tsegaye A. Integration opportunities for HIV and family planning services in Addis Ababa, Ethiopia: an organizational network analysis. BMC Health Serv Res. 2014;14:22. doi: 10.1186/1472-6963-14-22.

19. UNAIDS. Global AIDS update December 2002. Geneva: UNAIDS; 2002.

20. UNAIDS. Report on the global AIDS epidemic update 2002 [Internet]. Geneva: UNAIDS; 2002 [cited 2023 Jul 16]. Available from: https://data.unaids.org/pub/report/2002/brglobal_aids_report_en_pdf_red_en.pdf

21. Seckinelgin H. Who can help people with HIV/AIDS in Africa? Governance of HIV/AIDS and civil society. Voluntas. 2004;15(3):287–304.

22. Suseshna RP, et al. Community inclusion in PrEP demonstration projects: lessons for scaling up. Gates Open Res. 2019;3. doi: 10.12688/gatesopenres.13042.2.

23. Byanyima W, Lauterbach K, Kavanagh MM. Community pandemic response: the importance of action led by communities and the public sector. Lancet. 2023;40:253–5.

24. Provan KG, Milward HB. Do networks really work? A framework for evaluating public-sector organizational networks. Public Adm Rev. 2001;61(4):414–23. doi: 10.1111/0033-3352.00045.

25. Provan K, Veazie M, Staten L, Teufel-Shone N. The use of network analysis to strengthen community partnerships. Public Adm Rev. 2005;65(5):603–13. doi: 10.1111/j.1540-6210.2005.00487.x.

26. Nooraie RY, Khan S, Gutenberg J, Baker GR. A network analysis perspective to implementation: the example of health links to promote coordinated care. Eval Health Prof. 2019;42(4). doi: 10.1177/0163278718772887.

27. Worley C, Mirvis P. Studying networks and partnerships for sustainability: lessons learned. 2013. p. 261–91.

28. Valente T, Palinkas L, Czaja S, Chu K, Brown CH. Social network analysis for program implementation. PLoS One. 2015;10. doi: 10.1371/journal.pone.0131712.

29. Phillips GI, Lindeman P, Janulis P, Johnson AK, Beach LB, Stonehouse P, et al. Network analysis of organizations providing HIV services in Chicago: toward an integrated response to the HIV epidemic. J Public Health Manag Pract. 2022;28(2):143–51. doi: 10.1097/PHH.0000000000001165.

